# Similar and different: systematic investigation of proteogenomic variation between sexes and its relevance for human diseases

**DOI:** 10.1101/2024.02.16.24302936

**Authors:** Mine Koprulu, Eleanor Wheeler, Nicola D. Kerrison, Spiros Denaxas, Julia Carrasco-Zanini, Chloe M. Orkin, Harry Hemingway, Nicolas J. Wareham, Maik Pietzner, Claudia Langenberg

**Affiliations:** MRC Epidemiology Unit, University of Cambridge School of Clinical Medicine, Institute of Metabolic Science, Cambridge, CB2 0QQ, UK; Institute of Health Informatics, University College London, London, UK; Health Data Research UK, London, UK; British Heart Foundation Data Science Centre, London, UK; National Institute of Health Research University College London Hospitals Biomedical Research Centre; Computational Medicine, Berlin Institute of Health at Charité-Universitätsmedizin Berlin, 10117 Berlin, Germany; Precision Healthcare University Research Institute, Queen Mary University of London, London, UK; Blizard Institute and SHARE Collaborative, Queen Mary University of London, London, UK; Department of Infection and Immunity, Barts Health NHS Trust, London, E1 2AT, UK

## Abstract

To better understand sex differences in human health and disease, we conducted a systematic, large-scale investigation of sex differences in the genetic regulation of the plasma proteome (>5,000 targets), including their disease relevance.

Plasma levels of two-thirds of protein targets differed significantly by sex. In contrast, genetic effects on protein targets were remarkably similar, with very few protein quantitative loci (pQTLs, n=74) showing significant sex-differential effects (for 3.9% and 0.3% of protein targets from antibody- and aptamer-based platforms, respectively). Most of these 74 pQTLs represented directionally concordant effects significant in both sexes, with only 21 pQTLs showing evidence of sexual dimorphism, i.e. effects restricted to one sex (n=20) or with opposite directions between sexes (n=1 for CDH15). None of the sex-differential pQTLs translated into sex-differential disease risk.

Our results demonstrate strong similarity in the genetic regulation of the plasma proteome between sexes with important implications for genetically guided drug target discovery and validation.

## Main text

Many aspects of human development and health, including the age of onset, prevalence, and severity of many diseases differ between sexes (1–6), but the underlying mechanisms or extent to which genetic factors contribute to any differences remain largely unknown (7–9). Understanding genetically driven sex-differences at the molecular level, specifically proteins as the biologically active entity between the genome and the phenome, is important for basic and translational genetic research, including genetically anchored drug target discovery and validation.

Here, we examined the sex-specific genetic regulation of the plasma proteome across two cohorts and measurements from two different technologies. We contrast sex-differential protein abundance with sex-specific genetic regulation for 4,775 unique proteins, targeted by 4,979 unique aptamers in 4,403 females and 3,945 males (aged 29–64) from the Fenland study (10) and 1,463 unique proteins, targeted by 1,463 unique antibody assays among 25,904 females and 22,113 males (aged 49-60) from UK Biobank (11) (**Supplementary Table 1)** with 1,101 unique proteins being targeted by both platforms. We defined ‘female’ and ‘male’ sex by matching the recorded sex and sex chromosomes (XX for females and XY for males) for both studies. The recorded sex contained a mixture of self-reported sex and sex through medical records and it was not possible to distinguish sex from gender. We acknowledge the importance of distinguishing between sex and gender in research and that chromosomal make-up does not always align with self-identified gender.

Most protein targets (n=3,457 unique proteins out of 5,100 included in this study, 67.8%) showed significant sex differences (Bonferroni corrected p-value threshold for the number of assays measured by each technology, **see Methods**) in their plasma abundance in at least one cohort, including 521 (47.3%) overlapping targets with significant and directionally concordant effects (**Fig. 1**, **Supplementary Table 2**). Results exemplified large differences between the sexes, with a slightly larger number of protein targets showing higher levels in males compared to females across both technologies (**Fig. 1, Supplementary Table 2**). Adjustment for hormone replacement therapy/oral contraception or known sex-differential traits such as body mass index, low density lipoprotein cholesterol (LDL) levels, alanine transaminase (ALT) levels, smoking status and the frequency of alcohol consumption impacted only a moderate number of significant differences (9.34% and 18.63%, respectively; **Supplementary Table 2**). Proteins with the largest differences reflected sex-specific biology, e.g., specific expression in female- or male-specific tissues, reflected in plasma level differences of prostate-specific antigen (12) (PSA, beta [95% confidence interval (CI)]=1.56 [1.53-1.58], p=2.31×10^-2823^, UniProt: P07288), prokineticin 1 (beta [95% CI]= 1.25 [1.24 – 1.26], p=1.97×10^-7019^, UniProt: P58294), or follicle stimulating hormone (FSH, beta [95% CI]= −1.22 [−1.21 – −1.23], p=1.7×10^-6862^, UniProt: P01215), while others likely reflect the effect of sex-differences in body composition on plasma abundance of specific protein targets, such as leptin or adiponectin. We also observed strong sex-differences in established cardiovascular diagnostic markers such as NT-proBNP (beta [95% CI]= −0.78 [−0.74 – −0.82], p=3.33×10^-338^, UniProt: P16860) and troponin T (beta [95% CI]= 0.83 [0.79 – 0.87], p=9.86×10^-388^, UniProt: P45379).

**Figure 1:**
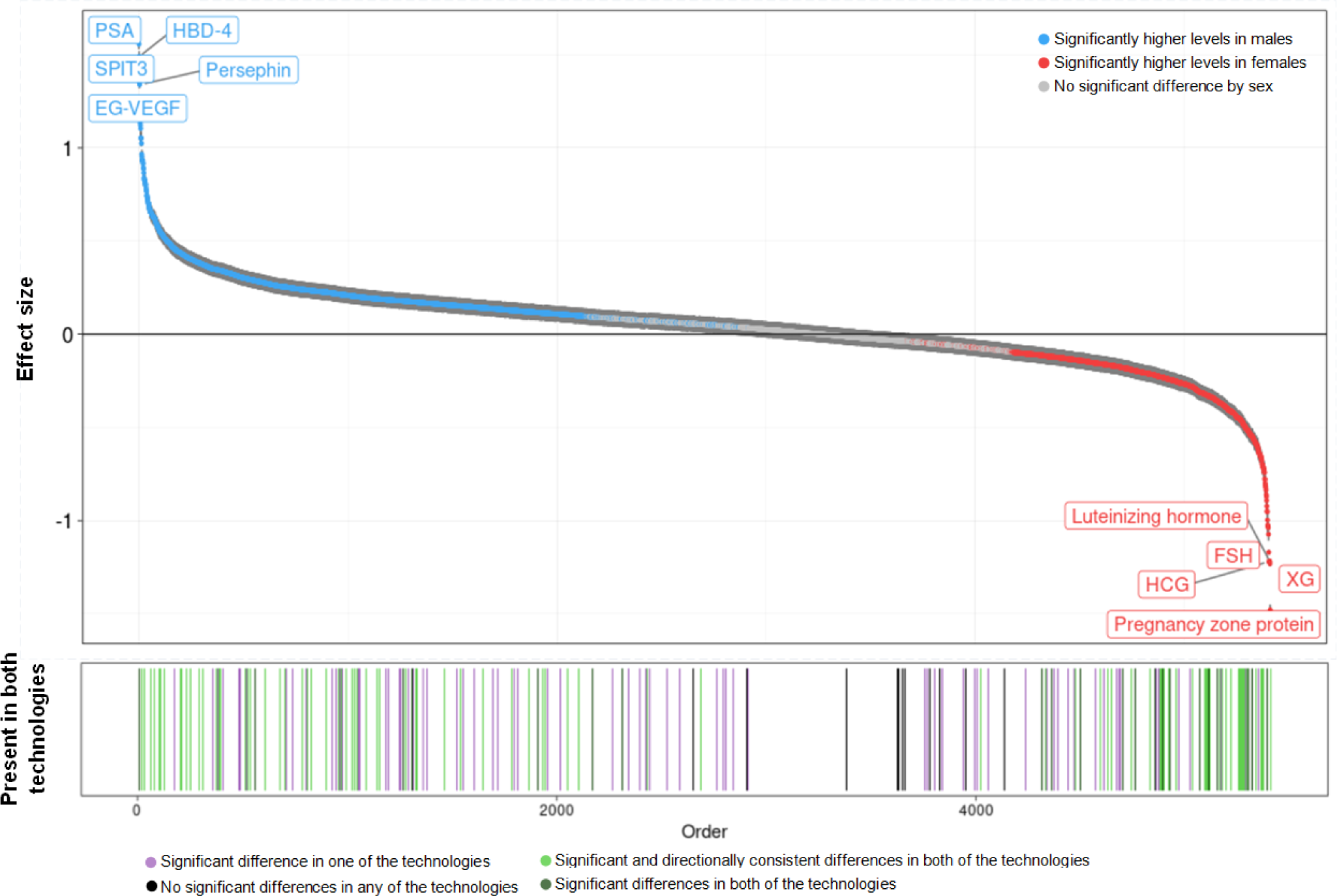
Sex differences in the abundance of 5,100 unique proteins measured by 4,979 unique aptamers and 1,463 unique antibody assays. The protein targets were ordered by their effect size in males. *Top panel*: The top panel shows the proteins for which the plasma abundance significantly differed by sex in at least one technology (p_het_<1.01×10^-5^ for aptamer-based and p_het_<3.42×10^-5^ for antibody-based technology were used as Bonferroni-corrected thresholds respectively). The proteins were coloured blue if they had significantly higher levels in males and red if they had higher levels in females. If the protein target was significant in both of the technologies, the effect size estimate from the more significant study was displayed. The dark grey vertical lines represent the 95% confidence intervals for the effect size estimates. *Bottom panel*: The bars in the bottom panel represent the proteins which were targeted by both aptamer-based and antibody-based platforms. The lines were coloured lighter green if the finding was significant and directionally consistent in both technologies, darker green if the finding was significant but not directionally consistent across technologies, lilac if the finding was only significant in one of the technologies and black if the finding was not significant in any of the technologies. Results can be found in Supplementary Table 2.

Males and females do not only differ by disease onset and severity, but also in drug response and a higher frequency of adverse drug reactions is observed in females compared to males (13). We identified a total of 92 proteins that are the targets of already approved drugs or drugs in early clinical trials (14) and showed significant differences between sexes in plasma abundance that were directionally consistent across cohorts. While plasma protein levels are not the primary target for most of those drugs, our results can potentially help understanding sex-differential drug effects.

We next performed sex-specific genome-proteome-wide association studies (15) to systematically identify sex-differential protein quantitative trait loci - ‘sd-pQTLs’ (**Supplementary Table 3 and 4)**. Despite the large number of pQTLs (p<5×10^-8^) identified in each sex (n_females_= 4,019, n_males_= 3,540 pQTLs aptamer-based and n_females_=13,013, n_males_=10,136 pQTLs for antibody-based technology (**Supplementary Figure 1)**), only very few pQTLs showed significant differences in effects between males and females (i.e. sex-differential effects), with 15 (p_het_<1.01×10^-11^) and 59 (p_het_<3.42×10^-11^) sd-pQTLs being identified for aptamer and antibody-based platforms, respectively (**Supplementary Table 3 and 4**). Most sd-pQTLs reside close to the cognate gene (cis-pQTLs; n=70.3%). We observed that the majority of sd-pQTLs (n=53 across technologies) showed effects that are directionally consistent and significant in both sexes but with significant, small effect size differences (i.e. same effect direction but different association strength between sexes). In other words, identified examples were predominantly sex differential rather than sex-dimorphic (i.e. only evident in one sex or different effect directions between sexes).

We observed no enrichment of sd-pQTLs on the X-chromosome or among druggable targets (p>0.05). We did not observe a clear bias towards protein encoding gene expression explicitly in reproductive tissues or breast for the proteins for which at least one cis or trans sd-pQTL was identified (16). Overall, 21 sd-pQTLs showed sex-dimorphic effects, with strong evidence of effect in one but not the other sex (p>5×10^-8^) for all, except for CDH15 where the sd-pQTL was significant in both males and females yet showed opposite effect directions. Some of the sex-dimorphic pQTLs mapped to proteins with established roles in only one of the sexes. For example, rs10843036 was specifically associated with pregnancy zone protein (PZP) in females (**Fig. 2, Supplementary Table 3**). Likewise, two protein targets with sd-pQTLs that were significant in males only (prostate and testis expressed protein 4 [PATE4, rs499684] and Kunitz-type protease inhibitor 3 [SPIT3, rs6032259]) have been reported to be involved in male fertility (**Fig. 2, Supplementary Table 3).** PATE4 has a reported function as a factor contributing to the copulatory plug formation in male fecundity in mouse models (17) and is predominantly expressed in prostate and testis (18). Similarly, SPIT3, encoded by *SPINT3*, is reported to be predominantly expressed in epididymis although Spint3 was reported to be dispensable for mouse fertility (19, 20). Although its sex-specific biological function is not clear, the male-specific sd-pQTLs for neural cell adhesion molecule 1 (NCAM-1) was replicated across both platforms (**Fig. 2, Supplementary Table 3 and 4)**.

**Figure 2:**
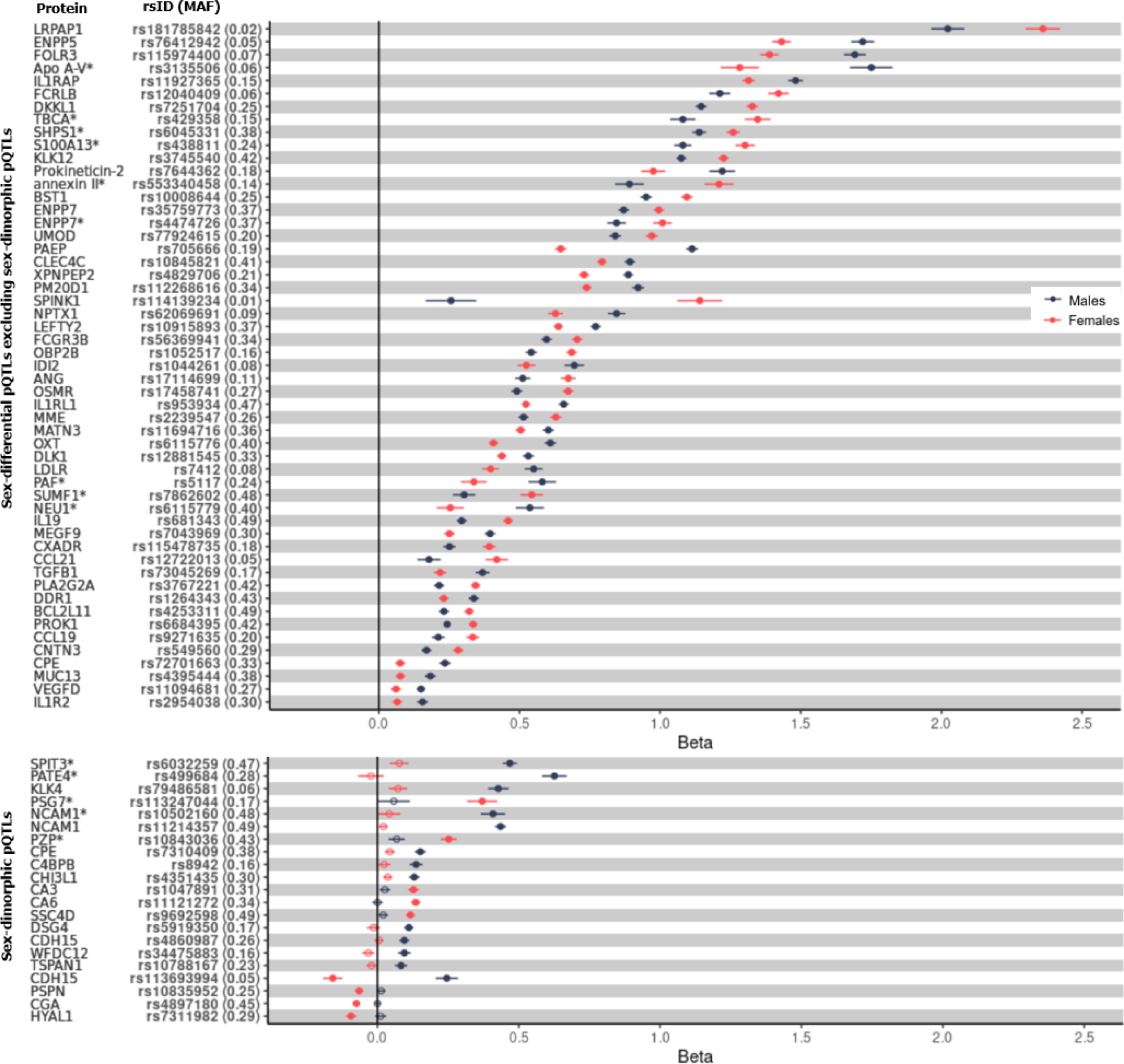
Forest plot of all identified sex-differential protein quantitative trait loci (sd-pQTLs) from both aptamer- (p_het_1.01×10^-11^) and antibody-based (p_het_3.42×10^-11^) technologies. The bottom panel presents sex-dimorphic pQTLs (not significant (p<5×10^-8^) in one sex or has opposing effect directions), whereas the top panel presents the remaining sex-differential pQTLs. The significant (p<5×10^-8^) pQTLs in each sex are represented by filled circles and non-significant ones are represented by hollow circles. Horizontal lines represent 95% confidence interval of each finding. Proteins with an asterisk(*) were measured using the aptamer-based technology, otherwise using antibody-based technology. Abbreviations: MAF, minor allele frequency.

For two proteins, we identified more than one sd-pQTL, further supporting a sex-differential or dimorphic genetic regulation (**Supplementary Table 4**). While the trans-sd-pQTL for cadherin −15 (CDH15) was male-specific, the cis-sd-pQTL (rs113693994, beta_females_[95% CI]= −0.16 [− 0.12 – −0.19], p_females_=9.8×10^-20^, beta_males_[95% CI]= 0.25 [0.21 – 0.28], p_males_=9.32×10^-35^) for CDH15 was the only example observed in this study where a pQTL was significant in both sexes but with opposite effect directions. It is therefore a pQTL that has not been identified in a sex-combined study (beta_sex_combined_[95% CI]= 0.02 [−0.01 – 0.04], p_sex_combined_=0.23). CDH15 acts a cell adhesion molecule that is involved in facilitating cell-cell adhesion and preserving tissue integrity and is highly expressed in brain and muscle. Similarly, carboxypeptidase E (CPE) which acts as an exopeptidase essential for the activation of peptide hormones (e.g. insulin) and neurotransmitters had both a cis and a trans sd-pQTL with both sd-pQTLs having stronger effects in males compared to females (**Supplementary Table 4**). Interestingly CPE has been implicated to have a role in osteoclast differentiation and CPE knockout mice displayed low bone mineral diversity and increased osteoclastic activity as well as being obese and displaying a diabetic phenotype (21, 22). However, neither CDH15 nor CPE have a clearly established sex-specific function or disease associations to date, although the fact that these two proteins have both cis and trans sex-specific genetic regulation might suggest their potential involvement in a sex-specific biological function.

We next attempted to identify any potential sex-differential phenotypic consequences of sd-pQTLs (n=74) across 365 diseases with more than 2,500 cases in UK Biobank. Despite 74 significant associations (Bonferroni corrected significance threshold of p<2.32×10^-6^ and p<9.31×10^-6^ for antibody- and aptamer-based technologies, respectively) between sd-pQTLs and disease risk in at least one sex, none of the associations showed evidence of sex-differential effects (**Supplementary Table 5 and 6**). This leaves the downstream physiological or pathological consequences of the identified sd-pQTLs yet to be determined (**Fig. 3)**.

**Figure 3:**
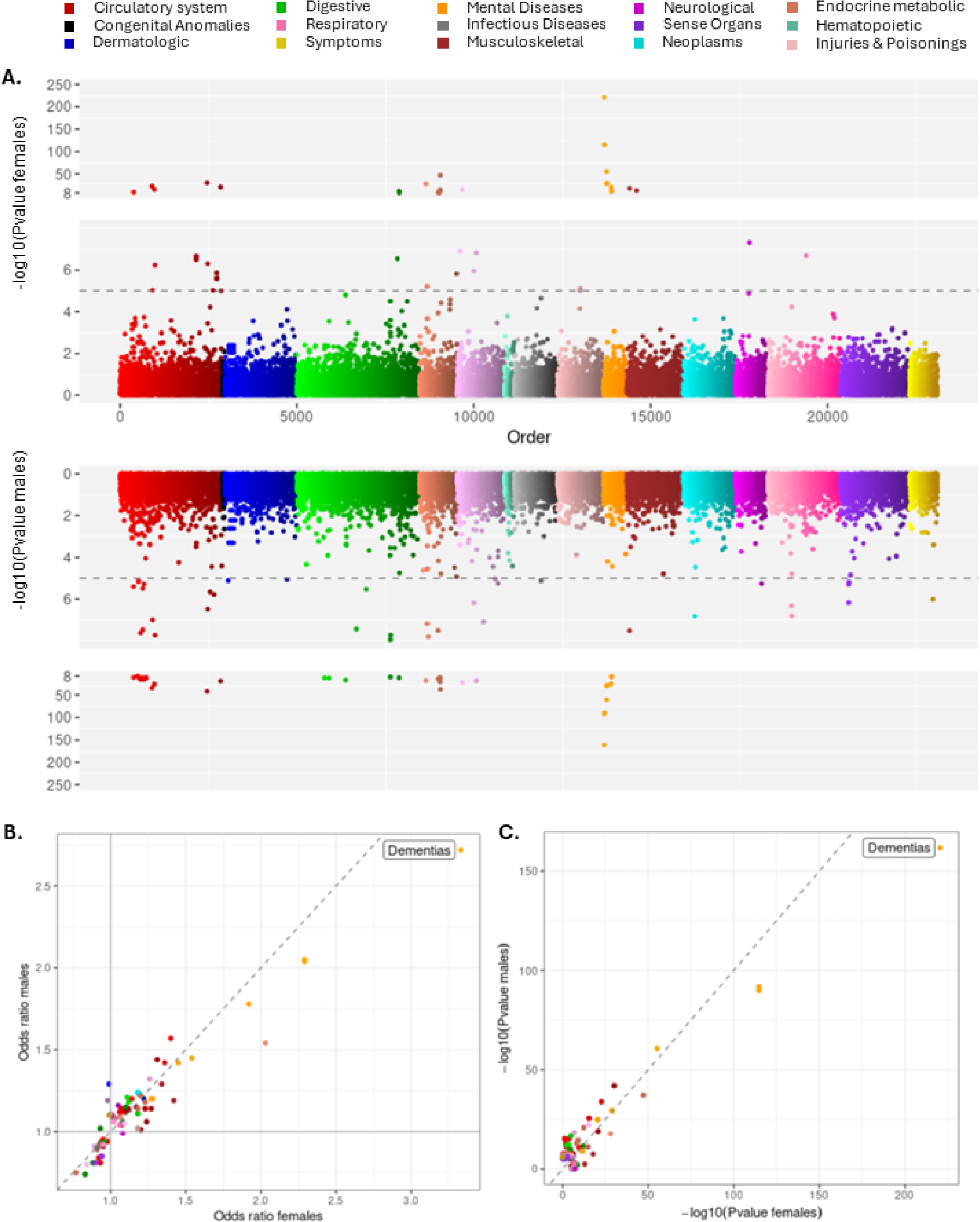
Phenotypic follow up of the identified sd-pQTLs with 365 disease outcomes with more than 2,500 cases in UK Biobank. **A. Miami plot of association of 74 sex-differential pQTLs (sd-pQTLs) with 365 disease outcomes among females on the top and among males at the bottom panel.** The disease categories have been coloured by disease categories. The horizontal dashed line represents a suggestive significance threshold of (p<1×10^-5^). **B. Comparison of odds ratios for the sd-pQTL – disease associations which meet the suggestive significance threshold (p<1×10^-5^) in males or females.** The diagonal dashed line represents the equality line (x=y). **C. Comparison of -log_10_(log_10_) transformed Pvalues for the sd-pQTL – disease associations which meet the suggestive significance threshold (p<1×10^-5^) in males or females.** The diagonal dashed line represents the equality line (x=y).

In summary, we identified sd-pQTLs for only a small proportion of the protein targets tested (0.3% and 3.9% of unique assays respectively for aptamer- and antibody-based technologies), a finding in line with previous sex-stratified analyses of tissue-specific gene-expression (16), and further confirmed that most sd-pQTLs act in a sex-differential rather than sex-dimorphic manner. This is in line with a recent study that reported trait variance difference between sexes can predominantly be explained by sex-differential ‘amplification effect’ (i.e. same effect direction yet different magnitudes of strength between sexes) (23).

Our study highlights two important conclusions. Firstly, the fact that we observe sd-pQTLs for a very small percentage of protein targets despite large differences in plasma protein levels emphasizes that the observed sex-differences are likely to be a result of intrinsic (e.g. hormone profiles) and extrinsic mechanisms (e.g. sex-differential lifestyle and risk-factor profiles) other than genetic regulation. This finding is in line with what has been reported for sex-differences observed for complex diseases, sex-differential genetic signals being identified for only a small proportion of common diseases (8). Secondly, our results suggest that the use of pQTLs in biomedical research, specifically for drug target discovery and causal inference will – with few exceptions - likely generate findings that are generalisable across sexes for the studied protein targets. However, larger studies should continue to evaluate sex differences as increased power could potentially uncover additional examples with biologically relevant sex-dimorphic effects.

Although only few, we did identify some sex-dimorphic genetic effects, with some reflecting sex-specific biology (e.g. PATE4, SPIT3, pregnancy zone protein) (24) acting possibly via steroid hormone responsive elements. Some of the other effects might possibly the result from differential environmental exposures between the sexes, as suggested for genetic variants affecting the risk for gout that may act through differential alcohol consumption as shown previously (25).

The restriction to proteins measured in plasma represents a notable limitation of our study, as sex-differential proteogenomic effects within tissues will unlikely be systematically reflected in plasma via secretion, natural cell turnover, or leakage. We obtained some evidence that larger sample sizes can identify a greater number of significant sd-pQTLs, but most act as weak modifiers of strong overall effects at protein encoding loci, and larger studies may possibly reveal even more subtle differences in regulation in trans for the previously targeted protein targets. Given the incomplete proteomic coverage (n=4,775 and n=1,463 unique proteins targeted by aptamer- and antibody-based platforms respectively, within a spectrum of over 20,000 proteins without taking post-translational modifications or different isoforms into account) as well as limited coverage of the genomic variant spectrum (i.e. rare variants or potentially ancestry-specific effects that we were not able to investigate), future studies might uncover new sd-pQTL signals as genomic and proteomic coverage continues to improve.

## Methods

### Study Participants

The Fenland study (26) is a population-based cohort of 12,435 participants of generally white-European ancestry, born between 1950 and 1975 who underwent detailed phenotyping at the baseline visit from 2005 to 2015. Participants were recruited from general practice surgeries in the Cambridgeshire region in the UK. The participants were excluded from the study if they were (i) clinically diagnosed with diabetes mellitus or a psychotic disorder, or (ii) pregnant or lactating, (iii) unable to walk unaided, or (iv) had a terminal illness. The study was approved by the Cambridge Local Research Ethics Committee (NRES Committee - East of England, Cambridge Central, ref. 04/Q0108/19) and all participants provided written informed consent.

This study used the largest subset of individuals from the Fenland study (**Supplementary Table 1**). 8,348 samples with both genotype information and proteomics measurements were taken forward for analyses after excluding ancestry outliers, related individuals or samples which have failed proteomics QC. The samples were well-balanced in terms of the participants from each sex: 4,403 (52.7%) females and 3,945 (47.3%) males were included in the study. Sex variable in Fenland study was based on general practitioners (GP) records. We only included participants with matching entries for the recorded sex and sex chromosomes (XX for females and XY for males). Individuals without matching entries were excluded from the study as a part of quality control as a mismatch can sometimes also be indicative of issues with genotyping protocol.

UK Biobank is a large-scale, population-based cohort with deep genetic and phenotypic data with the full cohort consisting of approximately 500,000 participants (11). The participants were recruited across centres in United Kingdom and were aged 40 to 69 years at the time of recruitment (11). Ethics approval for the UK Biobank study was obtained from the North West Centre for Research Ethics Committee (11/NW/0382) (11) and all participants provided informed consent. This study used the subset of European-ancestry individuals from UK Biobank where both genotype and proteomics measurements were available after excluding ancestry outliers or samples which have failed genomic or proteomics QC (n=48,017). 25,904 (53.9%) females and 22,113 (46.1%) males were included in the study (**Supplementary Table 1**). Sex in UK Biobank had two definitions, one was based on sex chromosomes (field 22001) and the other was contained a mixture of the sex the NHS had recorded for the participant and self-reported sex (field 31). We only included participants with matching entries for the recorded sex (from medical records or self-reported) and sex chromosomes (XX for females and XY for males).

### Genotyping and imputation

The Fenland-OMICS samples have been genotyped using the Affymetrix UK Biobank Axiom array. Sample-level and variant level QC criteria were applied as described elsewhere (26). In summary, the genotyped data was imputed to the HRC (r1) panel (27) using IMPUTE4 (https://jmarchini.org/software/) for the autosomes and Sanger Imputation Server for chromosome X (https://imputation.sanger.ac.uk/). The data was also imputed to the UK10K and 1000 Genomes Project 3 panels using and Sanger Imputation Server for both autosomes and chromosome X (28). Additional variants gained from the UK10Kp+1KGp3 imputation were added to the HRC imputed dataset. For basic quality control, variants were filtered for minor allele count (MAC) ≥3 using BCFtools (29) and INFO≥0.4 using QCTOOL v2.0.2 (https://www.well.ox.ac.uk/~gav/qctool_v2/) to eliminate variants with low imputation quality.

The UK Biobank samples were genotyped using the Affymetrix UK BiLEVE or the Affymetrix UK Biobank Axiom arrays. The following QC criteria was applied to the genotyping data (a) routine quality checks carried out during the process of sample retrieval, DNA extraction, and genotype calling; (b) checks and filters for genotype batch effects, plate effects, departures from Hardy Weinberg equilibrium, sex effects, array effects, and discordance across control replicates; and (c) individual and genetic variant call rate filters as previously described (11).

Genomic build GRCh37 was used throughout this study.

### Proteomic measurements

#### Aptamer-based platform

Fasting proteomic profiling of EDTA samples from Fenland study participants was performed by SomaLogic Inc. using the SOMAscan proteomic assay (v4). Relative protein abundances of 4,775 human protein targets were measured by 4,979 aptamers (SomaLogic V4). The quality control of the proteomic measurements has been described in detail previously (26). Briefly, hybridization control probes were used to generate a hybridization scale factor to account for variation in hybridization within runs. A ratio between each aptamer’s measured value and a reference value were computed to control for total signal differences between samples due to variation in overall protein concentration or technical factors. The median of these ratios was computed and applied to each dilution set (40%, 1% and 0.005%). Samples were removed if they were deemed by SomaLogic to have failed or did not meet our acceptance criteria of 0.25-4 for all scaling factors. In addition to passing SomaLogic QC, aptamers were filtered to only include human protein targets for subsequent analysis (n=4,979). Aptamers’ target annotation and mapping to UniProt (30) accession numbers as well as Entrez gene identifiers (31) were provided by SomaLogic and these were used those to determine genomic positions of protein encoding genes.

#### Antibody-based platform

The UK Biobank proteomic measurements were conducted by antibody-based Olink technology, Explore 1536 platform which uses Proximity Extension Assay (32). In summary, each protein is targeted by two unique antibodies with unique complimentary oligonucleotides, which only hybridize when they come into close proximity. This is subsequently quantified by next-generation sequencing. Normalized protein expression (NPX) units, which are reported on a log2 scale, are generated by normalization of the extension control and further normalization of the plate control. Further details about antibody-based proteomic measurements and QC have been described elsewhere, including the exclusion of samples due to poor quality and selective measurements with assay warnings (33).

### Sex-differences in protein abundances

We assessed the differential abundance levels of the 4,979 SomaLogic V4 aptamers between sexes in Fenland study. To estimate the effect of sex, a linear regression model was for implemented in R 3.6, using the inverse rank normalized proteomic values and including covariates age and test site in the model. A stringent Bonferroni-corrected threshold (corrected for n=4,979 aptamers; p<1.01×10^-5^) was applied.

1,463 protein targets from Olink Explore 1536 platform in UK Biobank were inverse rank normalized and subsequently restricted cubic splines function through ‘rsc’ function of ‘Hmisc’ package was applied to regress out technical covariates such as month of the blood draw, time that blood was drawn, fasting status and sample age in R v4.2.2. Similarly, the effect of sex assessed in abundance levels of the 1,463 protein targets from Olink Explore 1536 platform were assessed in a linear regression model using the inverse rank normalized residuals and including covariates age, age^2^ and proteomic batch in R v4.2.2. A stringent Bonferroni-corrected threshold (corrected for n=1,463 aptamers; p<3.42×10^-5^) was applied.

Sensitivity analyses was performed by including (a) participants who have undergone hormone replacement therapy or use oral contraception, or (b) for known sex-differential participant characteristics which were body mass index (BMI), low density lipoprotein (LDL) cholesterol levels, alanine transaminase (ALT) levels, smoking status and the frequency of alcohol consumption **(Supplementary Table 1**) as additional covariates in the analyses. The continuous variables (BMI, LDL and ALT) were inverse rank normalized before being included as covariates.

### Sex-stratified protein genome-wide association analysis (pGWASs)

For the aptamer-based platform, the protein abundances for 4,979 aptamers measured in Fenland study were inverse rank normalized and regressed for covariates age, test site and the first 10 genetic principal components in R v3.6. The residuals for each sex were used in the subsequent association analyses.

The fastGWA software (34) linear regression analysis was performed through GCTA version 1.93.2 for the sex-stratified genome-wide association analysis in each sex. Further variant level QC was also applied and only variants with MAC ≥3, INFO≥0.4, genotype missingness rate < 5% and MAF>1% were included in the downstream analyses.

For the antibody-based platform, the protein abundances for 1,463 assays measured in UK Biobank, the same residuals from the analyses of sex-differences in protein abundances (i.e. inverse rank normalized and technical covariates regressed out) were taken forward. Sex-stratified GWASs were performed using REGENIE v.3.4.1 (35) through performing two steps, as implemented by the software. In the first step, a whole-genome regression model is fitted for each phenotype to generate a covariate, which is subsequently included in the second step to allow for computationally-efficient analyses of a large number of phenotypes. For the first step, only high-quality variants passing the stringent QC criteria of MAF>1%, MAC>100, Hardy-Weinberg equilibrium p-value<1×10^-15^ and genotype missingness rate < 10% were used and SNPs were pruned for linkage-disequilibrium (LD), specified for 1000 variant windows, 100 sliding windows and r2<0.8 through Plink v.1.9. Subsequently, step 2 was applied to conduct sex-stratified the genome-wide association analyses for 1,463 protein targets with additional per-marker QC filters of MAC>50, MAF>1% and INFO>0.4.

### Heterogeneity analysis

We performed an inverse-variance fixed effects meta-analysis for each protein target using female-only and male-only summary statistics through METAL (v.2011-03-25) (34) to assess the heterogeneity in the genetic associations between sexes for each platform. We used a proteome and genome-wide Bonferroni corrected significance threshold (p_het_<1.01×10^-11^ and p_het_<3.42×10^-11^ respectively for aptamer- and antibody-based platforms) for heterogeneity p-value to define sex-differential protein quantitative trait loci (i.e. sd-pQTLs).

Significant genomic regions were defined by 1 Mb regions (±500 Kb on either side) around any variant with significant heterogeneity. The MHC region (chr6:25.5–34.0Mb) was treated as a single region. The regional sentinel variant for each genomic loci was defined as the most significant variant within the region. Variants were defined as cis-pQTLs if they were within the 1 Mb window (±500 Kb on either side) of the protein encoding gene and defined as trans-pQTLs if they were not within the 1 Mb window.

### Phenome-wide Association Study (PheWAS)

We have tested whether any of the significant sd-pQTLs showed heterogeneity between sexes in terms of their disease associations across the phenome. For this purpose, in each sex, we tested the association of sd-pQTLs with 365 binary diseases with more than 2,500 cases in UK Biobank. The binary disease categories were collated through clinical entities named ‘phecodes’ in UK Biobank, which were defined using the International Classification of Diseases, 10th Revision (ICD-10) and the International Classification of Diseases, 10th Revision, Clinical Modification (ICD-10-CM) codes from electronic health records, available in UK Biobank (36). We tested the association of each sd-pQTL with each phecode in each sex, using a logistic regression model in R v3.6 and adjusting for age, genotype batch, test centre, and the first ten genetic principal components in unrelated European participants. We have subsequently meta-analysed the female-only and male-only summary statistics using a fixed-effects meta-analyses through *metafor* package in R v3.6 to assess the heterogeneity of the association between sexes. To correct for multiple testing, p-value threshold for PheWAS was defined as p_het_< 9.31×10^-6^ and p_het_< 2.32×10^-6^ for aptamer- and antibody-based platforms respectively, which were corrected for the number of sd-pQTLs and number of phenotypes tested (n=365) in each platform.

## Acknowledgements

We are grateful to all Fenland volunteers and to the General Practitioners and practice staff for assistance with recruitment. We thank the Fenland Study Investigators, Fenland Study Co-ordination team and the Epidemiology Field, Data and Laboratory teams. SomaLogic proteomic measurements were supported and governed by a collaboration agreement between the University of Cambridge and SomaLogic.

The Fenland Study (DOI 10.22025/2017.10.101.00001) is funded by the Medical Research Council (MC_UU_12015/1). We further acknowledge support for genomics from the Medical Research Council (MC_PC_13046). This work is supported by the Medical Research Council (MC_UU_00006/1 - Etiology and Mechanisms) (C.L., E.W., M.P., N.K., and N.J.W.). M.K. is supported by Gates Cambridge Trust. H.H. is supported by Health Data Research UK and the NIHR University College London Hospitals Biomedical Research Centre. S.D. is supported by a) the BHF Data Science Centre led by HDR UK (grant SP/19/3/34678), b) BigData@Heart Consortium, funded by the Innovative Medicines Initiative-2 Joint Undertaking under grant agreement 116074, c) the NIHR Biomedical Research Centre at University College London Hospital NHS Trust (UCLH BRC), d) a BHF Accelerator Award (AA/18/6/24223), e) the CVD-COVID-UK/COVID-IMPACT consortium and f) the Multimorbidity Mechanism and Therapeutic Research Collaborative (MMTRC, grant number MR/V033867/1). J.C.Z. was supported by a 4-year Wellcome Trust PhD Studentship and the Cambridge Trust.

## Author contributions

M.K., M.P and C.L. designed the analysis and drafted the manuscript. M.K., E.W., S.D., N.K., J.C.Z, H.H. and M.P. have performed the quality control, data preparation or the bioinformatics analyses. N.J.W. is PI of the Fenland study. C.M.O. contributed to defining sex in this study and provided insights into broader concepts of sex and gender in research. All authors contributed to the interpretation of the results and critically reviewed the manuscript.

## Data availability

Data from the Fenland cohort can be requested by bona fide researchers for specified scientific purposes via the study website (www.mrc-epid.cam.ac.uk/research/studies/fenland/information-for-researchers/). Sex-stratified summary statistics will be made available upon publication.

Access to the UK Biobank genomic, proteomic and phenotype data is open to all approved health researchers (http://www.ukbiobank.ac.uk/). This research has been conducted using the UK Biobank resource under the application 44448.

