## Supplementary Materials for "Similar and different: systematic investigation of proteogenomic variation between sexes and its relevance for human diseases"

**Supplementary Figure 1: Summary of the study design and number of pQTLs identified.**

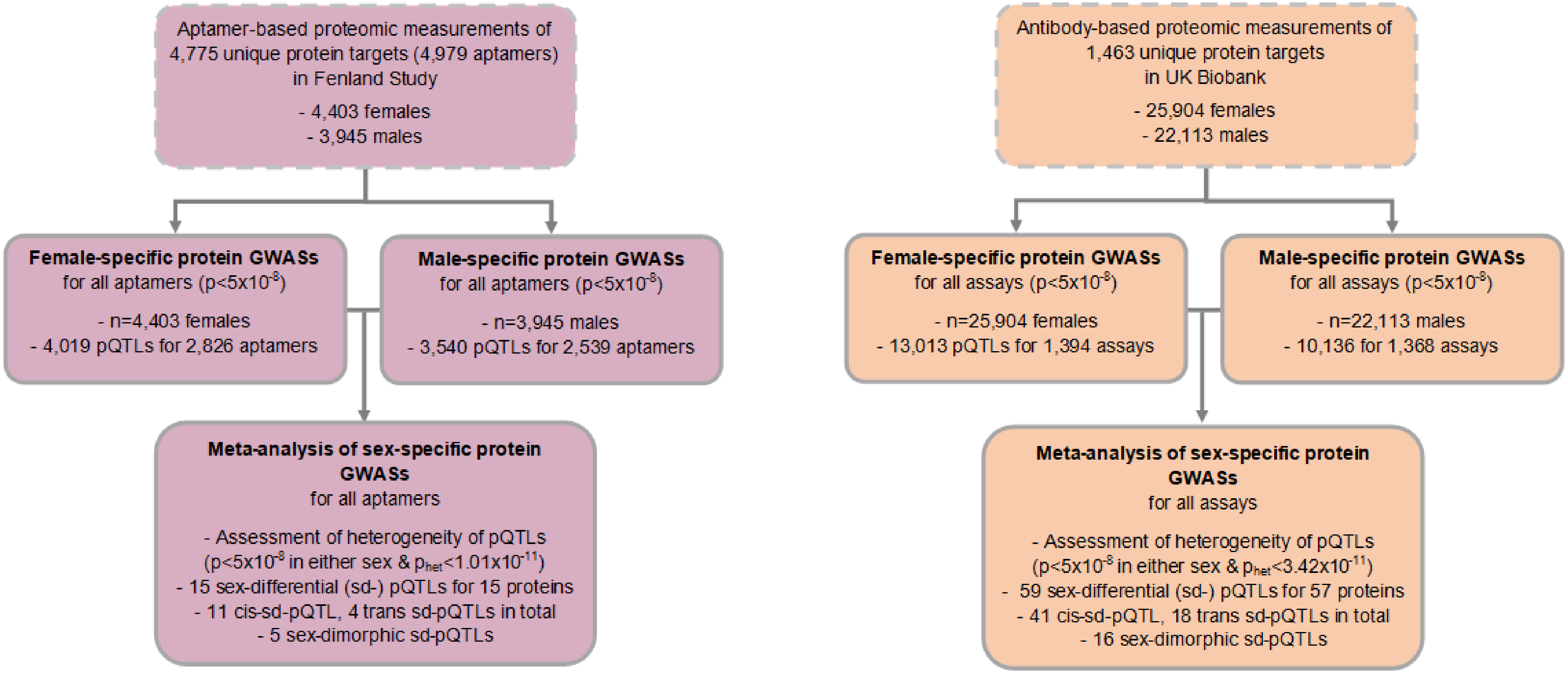

### Supplementary Tables

**Supplementary Table 1: Demographics of the Fenland and UK Biobank samples included in this study.**

**Supplementary Table 2. Observational sex-differences in protein abundances.** To estimate the effect of sex, a linear regression model was for implemented in R 3.6, including covariates age and test site in the model in each platform in the model. The columns with "sens_covars" suffix represent the results from sensitivity analyses where additional covariates (BMI, LDL, ALT, smoking status and alcohol consumption were included). The columns with "sens_HRT_OC" suffix represent the results from sensitivity analyses where use of hormone replacement therapy/oral contraception were included as an additional covariate. "SL" and "Olink" suffixes represents the results from aptamer- and antibody-based platform, respectively. Information on draggability based on common gene entries from Finan et al. (2017) (1).

**Supplementary Table 3. Sex-differences in the genetic regulation of protein abundances from aptamer-based technology in Fenland study**. This list includes all sd-pQTLs with a heterogeneity p-value<1.01e-11 between sexes from aptamer-based technology in Fenland study.

**Supplementary Table 4. Sex-differences in the genetic regulation of protein abundances from antibody-based technology in UK Biobank study.** This list includes all sd-pQTLs with a heterogeneity p-value<3.42e-11 between sexes from antibody-based technology in UK Biobank study.

**Supplementary Table 5. Phenome-wide association study results for 15 aptamer-based sd-pQTLs using 365 binary phenotypes with >2,500 cases in UK Biobank.**

**Supplementary Table 6. Phenome-wide association study results for 59 antibody-based sd-pQTLs using 365 binary phenotypes with >2,500 cases in UK Biobank.**
